# Trends and predictors of unmet need for family planning among women living with HIV in Zambia: implications for elimination of Mother to Child Transmission of HIV

**DOI:** 10.1101/2022.11.24.22282709

**Authors:** Edgar Arnold Lungu, Mwimba Chewe

## Abstract

**Introduction:** Prevention of Mother To Child Transmission (PMTCT) of HIV is one of the key strategies towards HIV epidemic control. Despite considerable progress in PMTCT of HIV over the past decade in Zambia, the country is yet to reach global and national target for elimination of Mother To Child Transmission of HIV. Avoidance of unintended pregnancy among women living with HIV provides is one of the cost-effective interventions in a comprehensive PMTCT of HIV approach. This study therefore aimed at ascertaining trends in and predictors of unmet need for family planning among women living with HIV in Zambia

**Methods:** The study employed a repeated cross sectional (RCS) study design, using data from the three (3) most recent consecutive rounds of the Zambia Demographic and Health Survey (ZDHS) conducted in 2007, 2013/2014 and 2018. The study used data from a total of 34,204 women aged 15-49 years from the three survey points, 2007, 2013/14 and 2018, among whom 4,985 were HIV positive, with a final sample size constituting 2,675 married women living with HIV. We used descriptive statistics and logistic regression analyses to respectively ascertain trends in and predictors of unmet need for family planning among married women living with HIV.

**Results:** Over the three survey points, unmet need for family planning among married women living with HIV has hardly declined, registering 22% in both the 2007 and 2018. Residence, age of women, household wealth, woman’s parity, employment, and age of spouse emerged as significant predictors of unmet need for family planning among women living with HIV in Zambia

**Conclusion:** Preventing one HIV infection in a child is averting lifetime costs of HIV treatment and associated healthcare costs. There is need to consider optimization of PMTCT interventions including shaping programming regarding prong 2 in a way that it responds to main causes of mother to child transmission of HIV in Zambia. Among other aspects, policy and practice needs to strengthen SRH/HIV integration and better target rural residents, younger women, those with high parity and consider positive male engagement to reduce unmet need for family planning among women living with HIV.

## Introduction

Prevention of mother-to-child transmission (PMTCT) of HIV has been integral to global HIV prevention interventions since the late 1990’s following the success of the short-course Zidovudine and single-dose nevirapine clinical trials (1-3). Subsequent recommendation from the World Health Organisation on the use of ARV drugs for PMTCT first issued in 2000 (4) also provided prominence to PMTCT of HIV programming. In the context of programming for Millennium Development Goals (MDGs), a more comprehensive four-pronged approach to PMTCT of HIV was launched by the United Nations in 2003 (5) and adopted by many countries with high HIV burden. The four-pronged approach focuses on provision of cost-effective interventions at critical stages during the perinatal period to prevent mother to child transmission of HIV (see figure 1).

**Figure 1:**
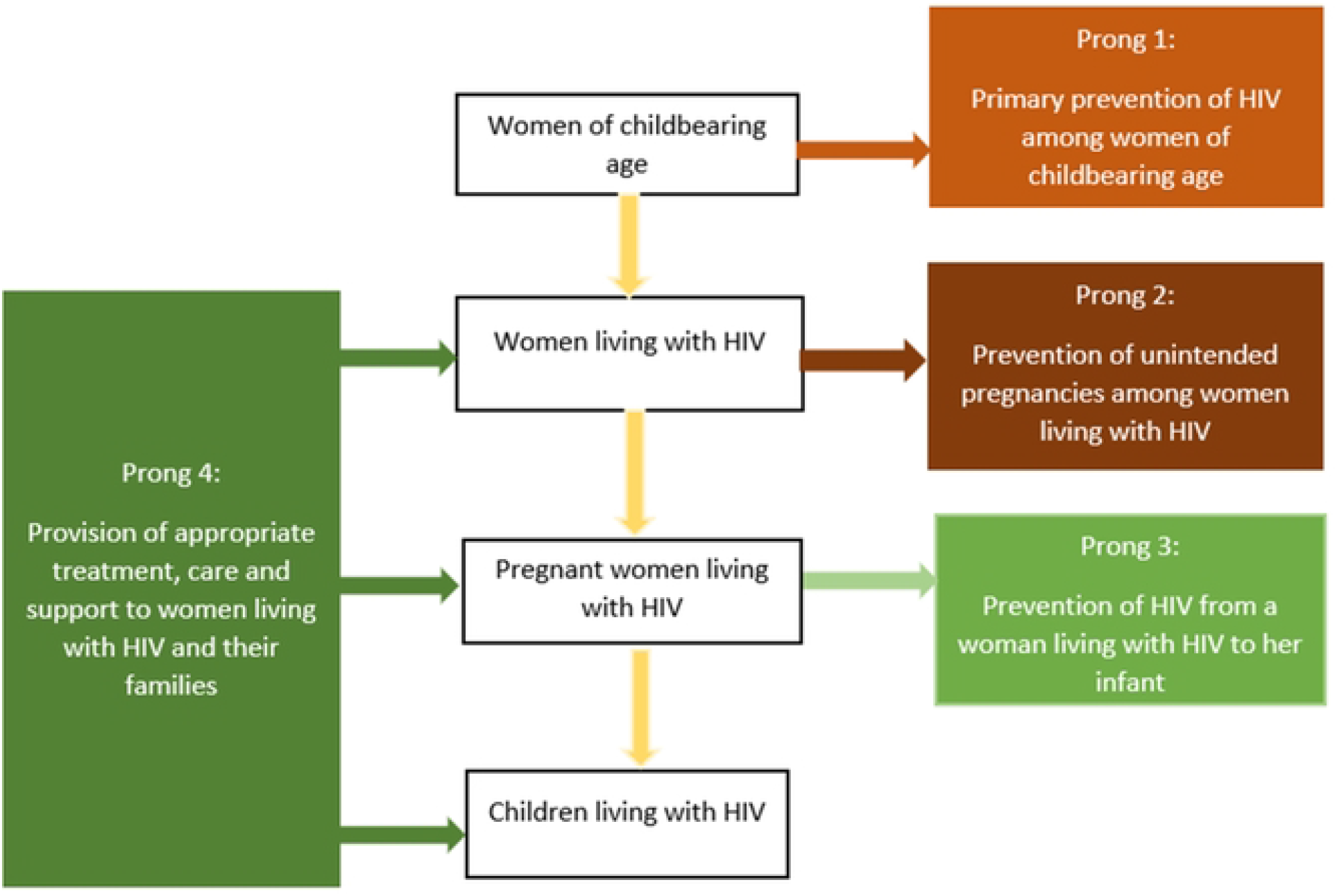
Four prongs for comprehensive prevention of mother to child transmission of HIV.

The first prong focusses on primary prevention of HIV among the general population, especially among women of reproductive age, given that averting new HIV infections in this population group entails parents that are free of HIV effectively eliminating the risk of transmission from parent to child. The second prong entails prevention of unwanted pregnancies among women living with HIV. Whilst more potent anti-retroviral (ARV) drugs currently exist and significantly reduce probability of mother to child transmission of HIV, avoidance of unintended pregnancy among women living with HIV provides a cost-effective way as it eliminates the risk of transmission. The third prong involves preventing mother-to-child transmission of HIV among pregnant and breastfeeding women who are living with HIV, and largely entails provision of HIV testing for case detection and linkage to sustained care and treatment especially provision of ARVs to reduce probability of mother to child transmission. The fourth prong requires provision of care, treatment and support services to women living with HIV, their children, and families with view to provide HIV prophylaxis for HIV exposed children, continued care and treatment for women to reduce new HIV infections in the breastfeeding period (5).

Although all four prongs are central to preventing mother to child transmission of HIV and promoting the health of women, PMTCT programmes in many countries have predominantly focused on identifying pregnant women with HIV and initiating them on treatment (prong 3) (6). Essentially, prong 3 and 4 interventions have been synonymous to PMTCT of HIV in many high HIV burden countries. Evidence on effectiveness of Option B+ and Treatment as Prevention (TasP) have arguably shaped the programming context in PMTCT (7) to date.

From 2015, UNAIDS advocated for a quarter for prevention aimed at revitalizing HIV prevention by calling for investing a quarter of resources in HIV and AIDS response to HIV prevention interventions (8) (which would predominantly serve prong 1 for PMTCT of HIV) given that ‘we cannot treat ourselves out of the HIV epidemic’. Whereas in theory, this made investment sense, actual implementation still principally focused on care and treatment interventions (mostly prongs 3 and 4). Granted this programming context, it is probable that the success of PMTCT programmes such as averting more than 110,000 new HIV infections among children between 2012 and 2021 in Zambia (9) and a 38 per cent reduction in MTCT of HIV between 2009 and 2014 (10) has largely been due to increasing coverage of prong 3 and 4 interventions.

Despite the prominence of prongs 3 and 4 in PMTCT programmes of high HIV burden countries including Zambia, evidence exists suggesting the centrality of prong 2 (prevention of unintended pregnancies among women living with HIV). For example, a study in PEPFAR supported countries estimated that annual number of unintended HIV-positive births averted by contraceptive use ranged from 178 in Guyana to over 120,000 in South Africa with minimum annual cost savings to prevent unwanted HIV-positive births reaching over 2.2 million United States Dollars in South Africa (11). Furthermore, a mathematical modelling study based on data from Uganda predicted that the effect of preventing unintended pregnancies in HIV positive women could contribute to PMTCT equally or more than ART (12). Noticeably, much of this evidence was in the pre-Option B plus and universal ART eligibility programming era (i.e. where ART was only given to the mother for up to 6 weeks and later up to two years postpartum for purposes of PMTCT of HIV).

Whereas, more recent estimates, in the context of high ART coverage among pregnant women show lower numbers of infant HIV infections averted by contraception than previously estimated, overarching conclusion from studies remains that contraception plays a significant role in preventing new infant HIV infections (13). On this basis, investing in prong 2 has significant merit to HIV epidemic control in general and specifically the elimination of Mother To Child Transmission (eMTCT) of HIV agenda. Arguably, the optimal benefits of investing in prong 2 have not been reaped (13) largely due to programming context. Expanding family planning services for women living with HIV presents opportunities to substantially contribute towards eMTCT of HIV in high HIV burden countries such as Zambia.

Zambia adopted and developed plans to pursue the global agenda for eMTCT of HIV with two impact targets, namely; an MTCT rate of HIV of less than 5% in breastfeeding population; and a population case rate of new pediatric HIV infections due to MTCT of equal to or less than 50 cases per 100,000 live births, (14 – 15). Whilst significant progress has been made, these targets were not achieved by 2021. Indeed, Zambia has sustained high coverage of HIV testing for pregnant women attending antenatal care which currently stands at 95% and consequently a high Anti-Retroviral Therapy (ART) coverage among pregnant women, currently standing at 97% in 2021 (9). Nonetheless, an MTCT of HIV rate at the end of exposure period (at the end of breast feeding) remains high at 8% falling short of the eMTCT of HIV targets. This reflects the need to re-examine current implementation including sustaining gains in prongs 3 and 4 while strengthening evidence and investment in prongs 1 and 2 which have hitherto been of relative low worth. Indeed, achieving the eMTCT agenda requires a comprehensive approach to PMTCT of HIV programming espousing optimal implementation of the four-pronged approach.

In the most recent Demographic and Health Survey (DHS) report, unmet need for family planning is estimated to be 19.7% in Zambia (16). There is paucity of consistent nationally representative data on unmet need for family planning among women living with HIV, with few studies reporting this statistic at a point in time (cross-sectional). In some cases, programme level data suggest an increasing uptake of family planning among women living with HIV in project’s lifespan (17). While there is some literature that has looked at unmet need for family planning among women in Zambia, there still remains a gap in literature on trends in unmet need for family planning amongst women living with HIV. Specifically, questions such as “ Has the unmet need for family planning among women living with HIV changed over time?”; “ What factors predict unmet need for family planning among women living with HIV in Zambia?” remain unanswered. Our study seeks to contribute to evidence in this regard and explore programmatic implications on the path to eMTCT of HIV in Zambia - a global priority country for HIV epidemic control and a recent global alliance on ending AIDS among children.

We argue that given centrality of prong 2 interventions in PMTCT of HIV and consequent HIV epidemic control, it is imperative to understand the trends and predictors of unmet need for family planning among women living with HIV. This is essential in designing interventions that would contribute to further reduction in MTCT of HIV.

## Methodology

### Study Design and Data sources

We undertook a secondary analysis of data from Demographic and Health Surveys (DHS) conducted in Zambia to understand trends and determinants of unmet need for family planning among women living with HIV. We adopted a repeated cross sectional (RCS) study design, using data from the three (3) most recent consecutive rounds of the Zambia Demographic and Health Survey (ZDHS). The RCS data structure is particularly beneficial because it introduces a dynamic component to the study of cross-sectional units and allows for the exploration of time-varying relationships which could broaden insight into the relationship being investigated (18)

Demographic and Health Surveys (DHS) are nationally representative population-based surveys conducted by the ICF Macro in collaboration with governments in about 90 countries, typically every four to five years. The DHS provide data on various health and demographic indicators including mortality, sexual and reproductive health, HIV, health status and health seeking, and child nutrition (19). In Zambia, five DHS have been conducted in 1992, 1996, 2001/02, 2007, 2013/2014 and 2018 (16).

Based on comparable data and study focus, our study used DHS data from surveys conducted in 2007, 2014 and 2018. The DHS use robust data quality assurance measures throughout the data collection and management process and the findings are highly regarded and used to inform policy and practice. We accessed all relevant DHS data sets upon request through the ICF Macro data management portal as custodians of DHS data. For each survey, two sets of data were availed, namely women’s data file constituting all sociodemographic and sexual and reproductive health data including family planning, and the HIV data file that included data on HIV testing and results.

### Definition of variables

In DHS, unmet need for family planning is typically defined to encompass two main groups, namely: (i) unmet need for spacing which means fecund women of childbearing age who would want to space their birth intervals but are currently not on any modern contraceptive method and (ii) unmet need for limiting referring to fecund women of childbearing age who would desire no additional children and are presently not on any modern contraceptive method (20).

Our outcome variable definition was consistent with the DHS variable definition albeit recognising that some discourse has contested and cited various challenges with the indicator definition. Some salient issues and challenges raised include, that: “ users of traditional methods are treated as non-users based on the implicit assumption that they lack access to or information concerning more effective alternatives; the concept of unmet need is based on the discrepancy between the future bearing wishes and contraceptive use rather than from a direct expression of need by respondents”, and that all estimates of unmet need are exclusively based on women’s reports (21). This notwithstanding, information on unmet need for family planning generated by DHS still reflects an acceptable measure and has been useful in informing policy and practice for many years.

To ascertain the relationship between different individual characteristics and unmet need for family planning among women living with HIV, we included several demographic and socioeconomic variables. The socioeconomic indicators we adopted included highest level of education, religion, whether respondent has worked in the past 12 months and the eligible women’s wealth level proxied by a wealth index. Demographic variables included age, partner’s age, and area of residence. Other variables included the total number of children ever born to a woman, partner’s education level, visit to health facility in the past year as well as whether the respondent is the primary decision maker on their health.

### Data Synthesis and Analysis

In the first stage of data synthesis, we merged two data sets (i.e. women’s HIV data set and family planning data set) from the ZDHS for the years 2007, 2013/14 and 2018 using the unique identification codes for the survey participants. This was done to accurately link the same participants from the two data sets to ensure that, although compiled separately, the two data sets were effectively linked to the extent that the family planning data and HIV data at individual level were indeed for the same woman.

Using the HIV test result data, we extracted married women who had a positive HIV test result as a subgroup of interest and were able to ascertain their status regarding family planning, specifically whether the need for family planning was met or unmet.

We calculated unmet need for family planning for women living with HIV using the standard DHS definition for each DHS data. We then undertook trend analysis using DHS surveys as data time points to observe any changes in unmet need for family planning among women living with HIV over time.

To determine the factors associated with unmet need for family planning amongst married women living with HIV in Zambia, we adopted a logistic regression analysis. We chose the logistic regression model because it is a widely used and accepted method of analysis for binary outcomes which is easy to execute and interpret (21) It also has notable advantages over the simple OLS regression analysis or Linear Probability Model such as better accuracy particularly with simpler datasets as well as a lower tendency towards over-fitting in non-complex datasets. Further, the multivariate logistic regression can be used to estimate probabilities and odds ratios while controlling for confounding variables.

## Results

### Sociodemographic characteristics

Our study included a total of 34,204 women aged 15-49 years that were representative of the three survey points, 2007, 2013/14 and 2018. Of the total women aged 15-49 years over the 3 survey periods, 4,985 (15%) were HIV positive, among whom 2,675 (53%) were married and formed the study sample. The HIV results are based on the robust HIV testing algorithm of the DHS. In respective DHS, the HIV prevalence ranged from 14% in 2018 to 16% in 2007.

Sociodemographic characteristics of our sample are as presented in table 1. On aggregate, respondents had predominantly attained primary education (48%), with the majority being urban residents (62%). Women aged 25-39 years accounted for about two thirds of the sample and close to two thirds were in the fourth (richer) and fifth (richest) wealth quintiles. The mean age of partners to women included in the sample was 40 (SD 0.24) and on average, women included in the study had about 4 children. The 2013/14 DHS period had the largest contribution to the overall sample

**Table 1:** Sociodemographic characteristics of participants.

| Variable | Married Women Living<br>with HIV<br>(N= 2,675) |  | Married Women Living<br>with HIV with met<br>need for FP<br>(N=1884) |  | Married Women Living<br>with HIV with un-met<br>need for FP<br>(N=556) |  |
| --- | --- | --- | --- | --- | --- | --- |
|  | Percent | n | Percent | n | Percent | n |
| **Residence** |  |  |  |  |  |  |
| Rural | 37.9% | 1,081 | 76% | 745 | 24% | 237 |
| Urban | 62.1% | 1,594 | 78% | 1,139 | 22% | 319 |
| **Age** |  |  |  |  |  |  |
| 15-19 | 2.0% | 56 | 71% | 40 | 29% | 16 |
| 20-24 | 11.3% | 316 | 83% | 258 | 17% | 53 |
| 25-29 | 21.0% | 568 | 81% | 442 | 19% | 105 |
| 30-34 | 23.7% | 610 | 77% | 449 | 23% | 132 |
| 35-39 | 20.3% | 536 | 76% | 378 | 24% | 117 |
| 40-44 | 14.2% | 383 | 72% | 235 | 28% | 91 |
| 45-49 | 7.4% | 206 | 67% | 82 | 33% | 42 |
| **Education** |  |  |  |  |  |  |
| No Education | 7.4% | 206 | 74% | 129 | 26% | 47 |
| Primary | 48.3% | 1,292 | 73% | 856 | 27% | 318 |
| Secondary | 38.7% | 1,016 | 82% | 773 | 18% | 170 |
| Higher | 5.7% | 161 | 87% | 126 | 13% | 21 |
| **Wealth Index** |  |  |  |  |  |  |
| Poorest | 8.8% | 254 | 73% | 169 | 27% | 63 |
| Poorer | 11.3% | 341 | 72% | 223 | 28% | 89 |
| Middle | 17.5% | 568 | 74% | 384 | 26% | 143 |
| Richer | 32.3% | 798 | 78% | 571 | 22% | 154 |
| Richest | 30.0% | 714 | 82% | 537 | 18% | 107 |
| **Religion** |  |  |  |  |  |  |
| Catholic | 16.6% | 448 | 81% | 329 | 19% | 80 |
| Protestant | 81.2% | 2,178 | 77% | 1,528 | 23% | 459 |
| Muslim | 0.6% | 16 | 55% | 9 | 45% | 6 |
| Other | 1.6% | 31 | 57% | 18 | 43% | 10 |
| **Year** |  |  |  |  |  |  |
| 2007 | 17.5% | 504 | 74% | 350 | 26% | 128 |
| 2013 | 45.8% | 1,255 | 79% | 914 | 21% | 244 |
| 2018 | 36.7% | 916 | 77% | 620 | 23% | 184 |
| **Currently Working** |  |  |  |  |  |  |
| No | 46.0% | 1,191 | 75% | 835 | 25% | 268 |
| Yes | 54.0% | 1,478 | 79% | 1,045 | 21% | 287 |
|  | **Mean** | **SD** | **Mean** | **SD** | **Mean** | **SD** |
| **Partners Age** | 39.73 | 0.24 | 38.59 | 0.25 | 41.06 | 0.61 |
| **Children Ever Born** | 3.56 | 0.05 | 3.4 | 0.10 | 4.20 | 0.05 |

Note: Data for some women living with HIV may have been excluded because some data elements were missing

### Levels and trends unmet need for contraceptives among married women living with HIV

Results show that over the three survey points, unmet need for family planning among married women living with HIV has marginally declined from 26% in 2007 to 21% in 2013/14 and increased slightly registering 23% in 2018 DHS (see table 1 and figure 1). Whilst there is a statistically significant difference between 2007 and 2013/18, the difference between 2018 and 2007 is not statistically significant (see table 2). Further analysis, disaggregating unmet needs for spacing and limiting reveal that the two categories contribute relatively the same to the overall unmet need for family planning with no survey point showing a difference of more than a percentage point.

**Figure 2:**
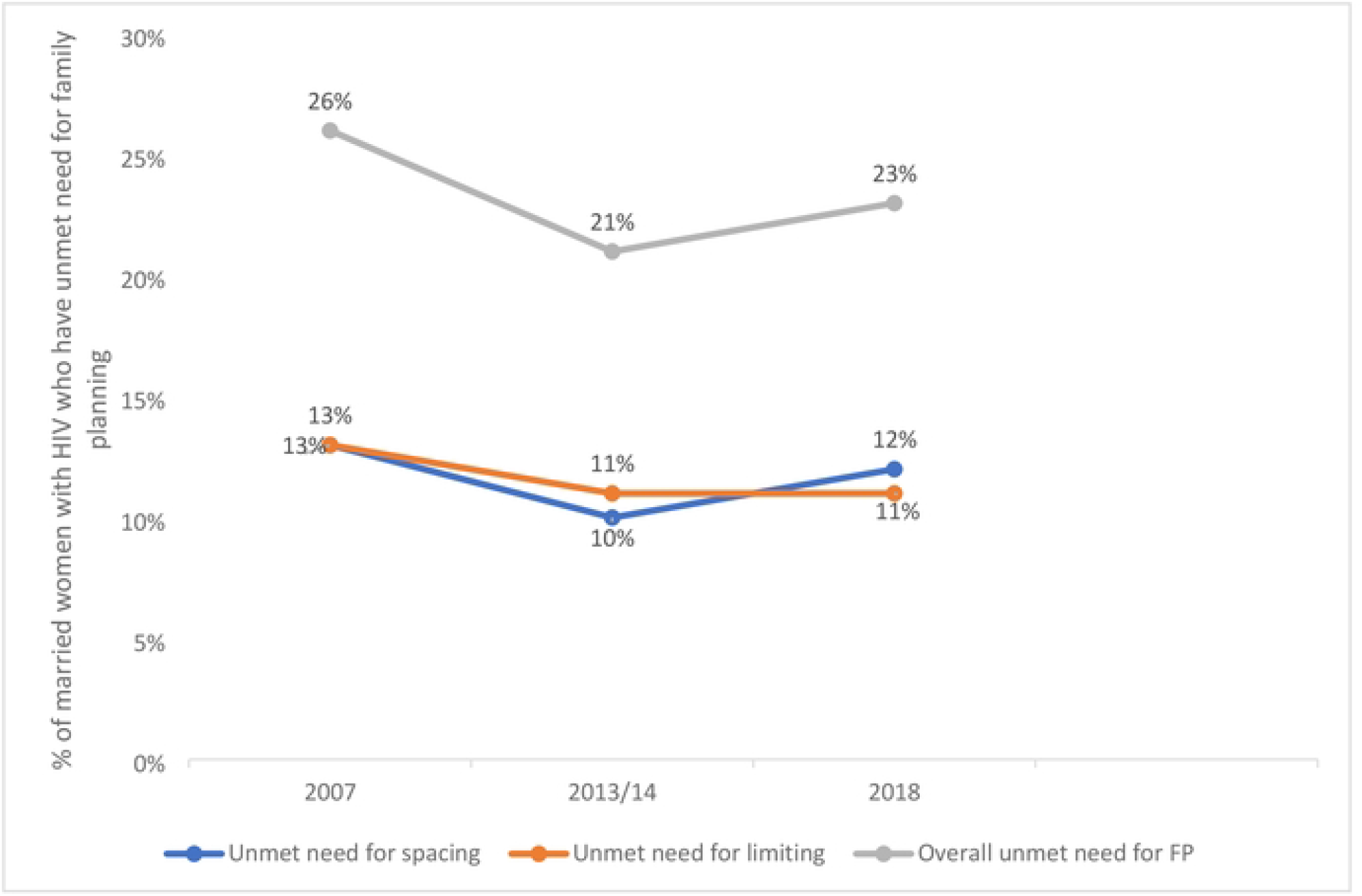
Trends in unmet need for family planning among married women living with HIV.

**Table 2:**
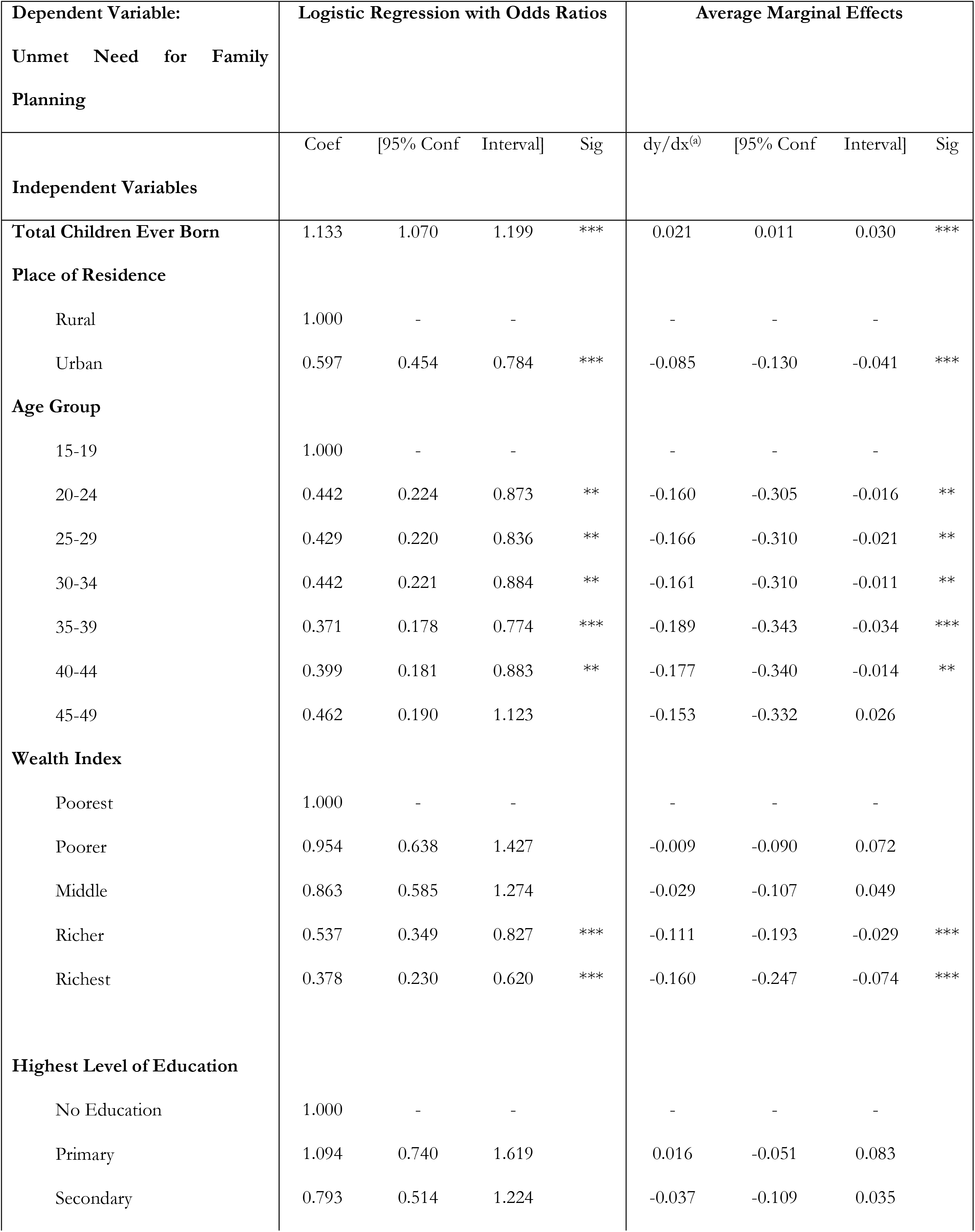
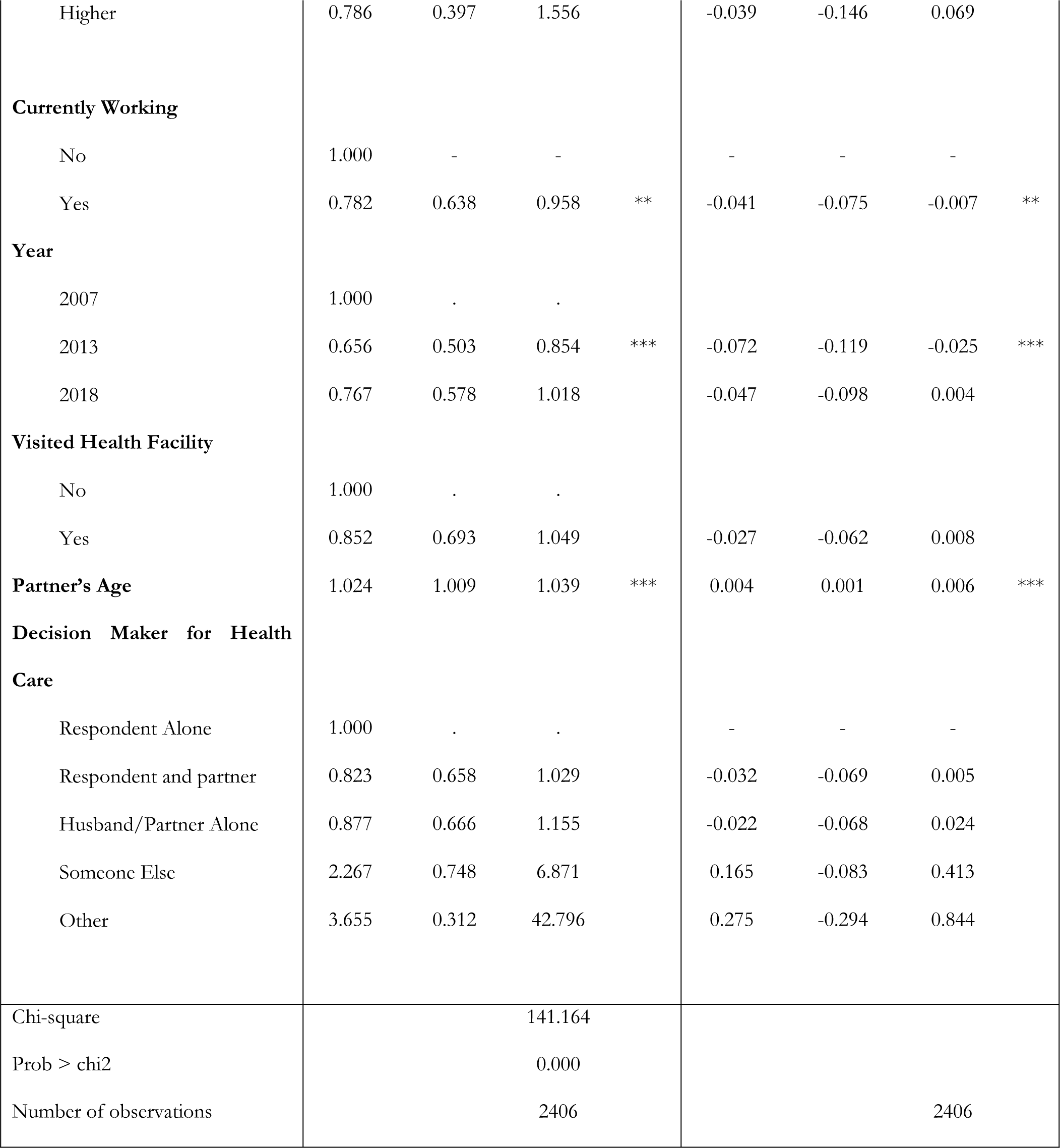
Logistic regression results of factors associated with unmet need for family planning among married women living with HIV.

### Factors associated with unmet need for family planning among married women living with HIV

Table 2 presents results of our multilevel logistic regression analysis for factors associated with unmet need for family planning among married women living with HIV over the three Zambia DHS survey points. Compared to women residing in rural areas, urban women are less likely to have unmet need for family planning (OR = 0.6095% CI: 0.45 - 0.78). Age of women emerged as a significant predictor of unmet need for family planning with women aged 20-44 years having lesser odds of having unmet need for family planning compared to adolescent girls aged 15-19 and the largest effect is observed in the 35-39 year old age group. There was no significant difference between women aged 45 years and above compared to adolescent girls aged 15-19 years.

Whilst there was no significant difference in unmet need for family planning among women in first two wealth quintiles (poorer and middle), compared to women in lowest (wealth quintile (poorest), those in fourth (richer) (OR = 0.54, 95% CI: 0.35 - 0.83) and fifth (richest) (OR = 0.39 95% CI: 0.23 - 0.62) wealth quintiles had lesser odds of having unmet need for family planning. The odds of having an unmet need for family planning increased with an increasing number of children ever born (parity) by their reproductive age at the point of the survey (OR = 1.13, 95% CI: 1.07– 1.19).

Women who were working at the time of the survey had less odds of having unmet need for family planning compared to women who reported not to be working (OR = 0.78, 95% CI: 0.64 - 0.96). Of the three survey periods included in this study, women living with HIV were less likely to have an unmet need for family planning during the 2013/14 survey period relative to 2007 albeit there was no significant difference between 2007 and 2018 survey points. Unmet need for family planning among women living with HIV increased with an increasing age of spouse/partner (OR = 1.02, 95% CI: 1.01 – 1.04).

Factors such as decision maker on health care, level of education and whether respondent visited health facility in 12 months preceding the survey, included in the regression model, were not significantly associated with unmet need for family planning.

## Discussion

The study findings suggest that unmet need for family planning among married women living with HIV marginally declined from 2007 to 2013/14 and thereafter increased again with no statistically significant difference between 2007 and 2018 survey levels. Essentially it has hardly changed in a sustained positive direction (i.e. reduction in unmet need) for slightly more than a decade – a period covered by this study. There is paucity of evidence showing trends in contraceptive use among women living with HIV in similar contexts making comparison with our study a difficult endeavour. Nonetheless, findings from some cross-sectional studies show similar levels reported in different survey points in our study. For example, a meta-analysis of unmet need for family planning among reproductive age women living with HIV in Ethiopia reported a pooled prevalence of 25% (22). Similarly, an analysis of DHS data from twelve African countries with an HIV prevalence of more than 3%, including Zambia, showed an unmet need for family planning among women with HIV ranging from 9% to 23% (23)

With strategies on reducing unwanted pregnancies among women living with HIV indicated as an important prong in PMTCT of HIV for the past two decades, stagnation of unmet need for family planning in this population group reflects some shortcomings that warrant attention if benefits of prong 2 are to be optimally realised. In contrast, between 2010 and 2020, anti-retroviral treatment coverage among pregnant women living with HIV (prong 3) in Zambia markedly increased from 58% to >95% (9) effectively narrowing the treatment gap.

As expected, an increasing trend in ART coverage among pregnant women living with HIV has corresponded with a declining trend in new HIV infections among children. There is also evidence that even modest decreases in the number of pregnancies to HIV-positive women could significantly reduce new HIV infections among children from both women living with HIV who are not on treatment (where prong 2 has the largest effect) and women living with HIV who are on treatment and desire to space or limit birthing (13). Moreover, some regional evidence suggests that an estimated “ 333,000 new infant infections could be averted annually, if all women in the sub-Sahara Africa who did not wish to become pregnant could have access to contraceptive services” (22). This evidence renders it highly probable that greater strides could have been made in reducing vertical transmission if unmet need for family planning reduced at a faster rate than current trends in Zambia.

Integrating family planning and ART services has emerged as one of the programming options to strengthen PMTCT prong 2 in many high HIV burden countries including Zambia. Multiple modalities such as; co-location of ART and family planning services, provider initiated family planning, capacity building of health workers for comprehensive SRH/HIV care (23 - 24) have been implemented with varying forms of design and results in different countries. In some cases this has yielded positive results with evidence from Ethiopia (25), Zambia (26) and Kenya (27)) suggesting increased contraceptive uptake and conversely reduction of unmet need for family planning where family planning and HIV services were integrated. Evidence of user satisfaction with services has also emerged from a systematic reviews (28). However, these promising practices have either been implemented at a smaller scale or have not been fully integrated in the healthcare service delivery system resulting in limited nationwide benefits and impacts. Furthermore, evidence is still sparse on long term benefits of some integration models (29). Consequently, unmet need for family planning has hardly declined among women living with HIV as per our study findings.

Current discourse in PMTCT of HIV has rightly tended to focus on more targeted interventions informed by evidence on main sources of new HIV infections among children along the PMTCT cascade of care. In Zambia, evidence shows that incident maternal HIV infections especially in the breastfeeding period account for majority of mother to child transmissions (9). In this epidemiological context for PMTCT, strengthening family planning services through integration with HIV services in the postpartum period, serves two integral functions for PMTCT of HIV.

Firstly, quality family planning counselling emphasizes dual protection from pregnancy and Sexually Transmitted Infections hence stands to directly contribute to reducing incident infections. Evidently, a study in Kenya found that integrating family planning and HIV services contributed to uptake of dual methods of protection from pregnancy and STI including HIV (27). Secondly, for women who still acquire HIV during the breastfeeding period, it offers a choice at an early stage of HIV infection, to prevent unwanted pregnancies subsequently, fully exploiting the benefits of prong 2.

Our analysis revealed that geographical residence (rural vs urban), age of woman, socioeconomic status, number of children ever born to a woman (parity), employment status and partner’s age constituted factors significantly associated with unmet need for family planning. These findings are largely consistent with other studies (22, 30-31) including a Zambian study which analysed unmet need for family planning using the 2018 DHS data (30). From a review of literature predictors of contraceptive uptake and unmet need for family planning are similar for HIV positive and HIV negative women.

Evidently, similar to our findings, studies in Tigray (32) and Dire Dawa (33) in Ethiopia, Nigeria and India (35) found that women living with HIV residing in rural areas were more likely to have unmet need for family planning than their counterparts from the urban areas. The probable explanation for this phenomenon is that women in rural areas have lower exposure to information and limited access to sexual and reproductive health services than their urban counterparts, as reported in surveys in Zambia and elsewhere (30, 31). Furthermore, in the Zambia country context, women in rural areas are arguably impacted more than their urban counterparts by other barriers to uptake of family planning services such as some deeply engrained cultural practices that propagate hesitancy to contraceptives and those that affect the autonomy of women to make reproductive choices.

Women in high socioeconomic status had lesser odds of having unmet need for family planning than their counterparts in lower socioeconomic strata. This renders support to similar findings reported in other studies in Ghana (36), Pakistan (37) and Ethiopia (38). The underlying explanation for this association is that women from wealthier households have greater ability to pay for direct and indirect costs associated with accessing modern contraceptive methods and are more likely to have better access to information on contraceptives and services than women from poorer households (39)

Consistent with other studies (23, 38, 40-41), our study found that women’s age is a significant predictor of unmet need for family planning. The pattern of association shows that women aged 20 to 44 years are less likely to have unmet need for family planning relative to younger women (below 20 years) although there is no statistically significant difference between older women (above 45 years) and younger ones (see table 2). This phenomenon may be explained by the comparator age group of adolescence (15-19 years) granted established evidence of multiple barriers to accessing family planning services for adolescent girls. These include stigma and discrimination, unfriendly attitude of health providers, inadequate information about contraceptives (42-44). However, the pathways explaining no difference between older and younger women are unclear albeit may be related to low-risk perception pertaining to unwanted pregnancy as reproductive age increases. It is worthwhile to note that another finding of this study suggests that unmet need for family planning among women living with HIV increased with an increasing age of spouse, a phenomenon that could be related to partner’s decision-making dynamics. Programmatic implications related to ongoing education and counselling on reproductive health choices throughout the reproductive period are therefore apparent.

Our study findings also suggest that the more children a woman has the higher the chances they have an unmet need for family planning. This is in line with study findings from Zambia (30) Ethiopia (45-46) and Kenya (47) including a multi-country study that analysed nineteen DHS survey data from Sub-Sahara Africa (48). It is probable that with higher parity, women will have attained the desired number of children hence a greater need for limiting and in the context of limited access to contraceptive services the unmet need is potentially high. Consequently, a programming implication entails the need for targeted family planning education and counselling for women with higher parity.

Our study confirmed findings of earlier studies (37) where employed women had less odds of unmet need for family planning. The causal pathways for this phenomenon relate to the potential for employed women to be more empowered, have the financial capacity, better exposure to information to make reproductive health choices including uptake of family planning services.

While education has been shown to be a significant predictor of contraceptive uptake, and conversely of unmet need for family planning among women in other studies (33-34, 49), our findings did not demonstrate any association. It is however consistent with a study in Zambia that used the 2018 DHS data and involved all married women (not merely those with an HIV positive result) (30). The mechanism for this phenomenon (lack of association with education) is unclear although it points to the importance of emphasizing family planning specific education and counselling irrespective of a woman’s education level.

Our study strengths relate to the use of data from DHS which are large population-based surveys considered robust enough to be nationally representative and frequently used to inform policy and practice. One salient limitation is that, while trends from cross-sectional survey points are useful and in our study context, most appropriate, they are not as robust as longitudinal studies for which temporal relationship should have been better defined. Other limitations common to surveys including recall and reporting bias are also applicable to our study although we acknowledge the rigor of DHS in minimizing these biases.

## Conclusion

Our study findings show that unmet need for family planning among women living with HIV have stagnated for more than a decade despite a long-standing programming context and evidence on the need to improve contraceptive uptake for women living with HIV not desiring to have children as part of PMTCT of HIV and improving maternal health.

Preventing one HIV infection in a child is averting lifetime costs of HIV treatment and associated healthcare costs. Granted that advancing PMTCT prong 2 is evidence based, it is imperative that significant efforts are invested in reducing unmet need for contraceptives among women living with HIV. The findings of this study point to various other implications for policy and practice. There is need to consider optimization of PMTCT interventions including shaping programming regarding prong 2 in a way that it responds to main causes of mother to child transmissions in Zambia. Strengthening and scaling up integration of SRHR/HIV and SGBV services including family planning and HIV prevention, care and treatment programme remains an option for which evidence of effectiveness and examples for implementation modalities already exist. The extent to which Zambia pursues the last mile towards path to elimination of mother to child transmission of HIV partly depends on optimization of proven cost-effective interventions. Reducing unmet need for contraceptives among women living with HIV continues to hold promise in this regard.

## Data Availability

Data used for this work is publicly available, on request, through the DHS Program

## Acknowledgements

The authors acknowledge the DHS Program for allowing access to the DHS data sets for Zambia which enabled us to undertake analysis and produce this manuscript.

## Supporting Information

S1 Fig 1: Four prongs for comprehensive prevention of mother to child transmission of HIV

S2 Fig 2: Trends in unmet need for family planning among married women living with HIV

## References

1. Sperling RS, Shapiro DE, Coombs RW, Todd JA, Herman SA, McSherry GD, et al. Maternal viral load, zidovudine treatment, and the risk of transmission of human immunodeficiency virus type from mother to infant. Pediatric AIDS Clinical Trials Group Protocol 076 Study Group. New England Journal Med. 1996; 335(22): 1621-9. 3.

2. Guay LA, Musoke P, Fleming T, Bagenda D, Allen M, Nakabiito C, et al. Intrapartum and neonatal singledose nevirapine compared with zidovudine for prevention of mother-to-child transmission of HIV1 in Kampala, Uganda: HIVNET 012 randomised trial. Lancet. 1999; 354(9181): 795–802.

3. Shaffer N, Chuachoowong R, Mock PA, Bhadrakom C, Siriwasin W, Young NL, et al. Short-course zidovudine for perinatal HIV-1 transmission in Bangkok, Thailand: a randomised controlled trial. Bangkok Collaborative Perinatal HIV Transmission Study Group. Lancet. 1999; 353(9155)

4. WHO. Prevention of Mother-To-Child Transmission (PMTCT) Briefing Note October 1st, 2007. Available from: https://www.who.int/hiv/pub/toolkits/PMTCT%20HIV%20Dept%20brief%20Oct%2007.pdf

5. Interagency Task Team (IATT) on Prevention of HIV Infection in Pregnant Women, Mothers and their Children. Guidance on global scale-up of the prevention of mother to child transmission of HIV: towards universal access for women, infants, and young children and eliminating HIV and AIDS among children. 2007. Available from: https://apps.who.int/iris/handle/10665/43728.

6. Hairston, A.F., Bobrow, E.A., & Pitter, C.S. (2012) Towards the Elimination of Pediatric HIV: Enhancing Maternal, Sexual, and Reproductive Health Services. International Journal of Maternal and Child Health and AIDS: 2012; 1(1): 6–16.

7. WHO. Antiretroviral treatment as prevention (TasP) of HIV and TB. 2012. Available from: https://apps.who.int/iris/bitstream/handle/10665/70904/WHO_HIV_2012.12_eng.pdf?sequence=1

8. UNAIDS. Invest in HIV Prevention: quarter for HIV prevention. Available from: https://www.unaids.org/sites/default/files/media_asset/JC2791_invest-in-HIV-prevention_en.pdf

9. UNAIDS. 2021. Available from: https://aidsinfo.unaids.org/

10. UNAIDS and PEPFAR. 2015 progress on the global plan towards the elimination of new HIV infections among children and keeping their mothers alive. 2015. Available from: https://www.unaids.org/sites/default/files/media_asset/JC2774_2015ProgressReport_GlobalPlan_en.pdf.

11. Reynolds HW, Janowitz B, Wilcher R, Cates W. Contraception to prevent HIV-positive births: current contribution and potential cost savings in PEPFAR countries. BMJ Journals. 2008; 84 (2) 49–53. doi: 10.1136/sti.2008.030049

12. Hladik W, Stover J, Esiru G, Harper M, Tappero J. The Contribution of family planning towards the prevention of vertical HIV transmission in Uganda. PLoS One. 2009; 4(11): doi: 10.1371/journal.pone.0007691

13. Sherwood J, Lankiewicz E, Roose-Snyder B, Cooper B, Jones A, Honermann B. The role of contraception in preventing HIV-positive births: global estimates and projections. BMC Public Health. 2021; 21:536 https://doi.org/10.1186/s12889-021-10570-w

14. Ministry of Health. National plan for the elimination of Mother To Child Transmission of HIV and syphilis. Lusaka, Zambia.

15. WHO. Global guidance on criteria and processes for validation: elimination of mother-to-child transmission of HIV, syphilis and Hepatitis B virus. Geneva: World Health Organization; 2021.

16. The DHS Program. Zambia Demographic and Health Survey 2018

17. Hancock NL, Chibwesha CJ, Bosomprah S, Newman J, Mubiana-Mbewe M, Sitali ES, Chi BH. Contraceptive use among HIV-infected women and men receiving antiretroviral therapy in Lusaka, Zambia: a cross-sectional survey. BMC Public Health. 2016;16(1):392

18. Matthew J, Lebo Weber W. An effective approach to the repeated cross-sectional design. American Journal of Political Science. 59(1) DOI:10.1111/ajps.12095

19. The DHS Program. Available from: http://dhsprogram.com/What-We-Do/Survey-Types/DHS.cfm.

20. MEASURE DHS. Family planning and Reproductive health indicators database: Unmet need for family planning. Available from: https://www.measureevaluation.org/prh/rh_indicators/family-planning/fp/unmet-need-for-family-planning

21. Hosmer DW, Hosmer T, Cessie SLE, Lemeshow S. A comparison of goodness-of-fit tests for the logistic regression model. 1997; 1996:965–80.

22. Kefale B, Adane B, Damtie Y, Arefaynie M, Yalew M, Andargie A. Unmet need for family planning among reproductive-age women living with HIV in Ethiopia: a systematic review and meta-analysis. PLoS ONE. 2021; 16(8): e0255566. https://doi.org/10.1371/journal.pone.0255566

23. MacQuarrie KLD. Unmet need for family planning among young women: levels and trends. DHS Comparative Reports No. 34. Rockville, Maryland, USA: ICF International; 2014. 58.

24. Nkhoma L, Sitali DC, Zulu JM. Integration of family planning into HIV services: a systematic review. Annals of Medicine. 2022; 54:1, 393-403, DOI: https://doi.org/10.1080/07853890.2021.2020893

25. Demissie DB, Mmusi-Phetoe R. Integration of family planning services with HIV treatment for women of reproductive age attending ART clinic in Oromia regional state, Ethiopia. Reprod Health. 2021; 18, 102:. https://doi.org/10.1186/s12978-021-01157-0

26. Hancock NL, Vwalika B, Sitali ES, Mbwili-Muleya C, Chi BH, Stuart GS. Evaluation of service quality in family planning clinics in Lusaka, Zambia. Contraception. 2015;92(4): 345–9. doi: 10.1016/j.contraception.2015.06.025

27. Chen Y, Begnel E, Muthigani W, Achwoka D, Mcgrath CJ, Singa B, Gondi J, Ng’ang’a L, Langat A, John-Stewart G, Kinuthia J, Drake AL. Higher contraceptive uptake in HIV treatment centers offering integrated family planning services: a national survey in Kenya. Contraception. 2020;102(1): 39–45. doi: 10.1016/j.contraception.2020.04.003.

28. Narasimhan M, Yeh PT, Haberlen S. et al. Integration of HIV testing services into family planning services: a systematic review. Reprod Health. 16 (1), 61 (2019). https://doi.org/10.1186/s12978-019-0714-9

29. Lindegren ML, Kennedy CE, Bain-Brickley D, Azman H, Creanga AA, Butler LM, Spaulding AB, Horvath T, Kennedy GE. Integration of HIV/AIDS services with maternal, neonatal and child health, nutrition, and family planning services. Cochrane Database of Systematic Reviews. 2012; 9. DOI: 10.1002/14651858.CD010119.

30. Namukoko H, Likwa R.N, Hamoonga TE, Phiri M. Unmet need for family planning among married women in Zambia: lessons from the 2018 Demographic and Health Survey. BMC Women’s Health. 2022; 22, 137. https://doi.org/10.1186/s12905-022-01709-x

31. Anik AI, Islam MR, Rahman MS. Association between socioeconomic factors and unmet need for modern contraception among the young married women: A comparative study across the low- and lower-middle-income countries of Asia and Sub-Saharan Africa. PLOS Glob Public Health. 2022; 2(7): e0000731. https://doi.org/10.1371/journal.pgph.0000731

32. Melaku YA, Zeleke EG. Contraceptive utilization and associated factors among HIV positive women on chronic follow up care in Tigray region, Northern Ethiopia: A cross sectional study. PLoS One. 2014; 9 (4):1–10. https://doi.org/10.1371/journal.pone.0094682 PMID: 24743241

33. Dejene H, Abera M, Tadele A. Unmet need for family planning and associated factors among married women attending antiretroviral treatment clinics in Dire Dawa City, Eastern Ethiopia. PLoS ONE. 2021; 16(4): e0250297.https://doi.org/10.1371/journal.pone.0250297

34. Fagbamigbe AF, Afolabi RF, Idemudia ES. Demand and unmet needs of contraception among sexually active in-union women in Nigeria: Distribution, associated characteristics, barriers, and program implications. SAGE Open. 2018; 8(1). https://doi.org/10.1177/2158244017753046 PMID: 30221033

35. Koshewara P, Fuke RP. Identifying the Unmet need of contraception among HIV seropositive women attending Antiretroviral Treatment (ART) clinic in tertiary care centre. J Evid Based Med Healthcare. 2019; 6(11):893–8

36. Lauro D. Abortion and contraceptive use in sub-Saharan Africa: how women plan their families. Afr J Reprod Health/La Revue Africaine De La Santé Reproductive. 2011; 15(1):13–23

37. Asif MF, Pervaiz Z. Socio-demographic determinants of unmet need for family planning among married women in Pakistan. BMC Public Health. 2019;19(1):1–8.

38. Tadele A, Abebaw D, Ali R. Predictors of unmet need for family planning among all women of reproductive age in Ethiopia. Contracept Reprod Med. 2019; 4(1):6

39. Ahinkorah BO, Ameyaw EK, Seidu AA, Ahinkorah, et al. Socio-economic and demographic predictors of unmet need for contraception among young women in sub-Saharan Africa: evidence from cross-sectional surveys. Reprod Health. 2020: 17:163 https://doi.org/10.1186/s12978-020-01018-2

40. Juarez F, Gayet C, Mejia-Pailles G. Factors associated with unmet need for contraception in Mexico: evidence from the National Survey of demo-graphic dynamics 2014. BMC Public Health. 2018;18 (1):546.

41. Dejenu G, Ayichiluhm M, Abajobir AA. Prevalence and associated factors of unmet need for family planning among married women in Enemay District, Northwest Ethiopia: a comparative cross-sectional study. J Community Med Health Educ. 2013; 13:4.

42. Bain EL, Amu H, Tarkang EE. Barriers and motivators of contraceptive use among young people in Sub-Saharan Africa: a systematic review of qualitative studies. PLoS ONE. 2021; 16(6): e0252745. https://doi.org/10.1371/journal.pone.0252745

43. Ochako R, Mbondo M, Aloo S, Kaimenyi S, Thompson R, Temmerman M, et al. Barriers to modern contraceptive methods uptake among young women in Kenya: a qualitative study. BMC public health. 2015; 15(1):118. https://doi.org/10.1186/s12889-015-1483-1 PMID: 2588467525.

44. Hokororo A, Kihunrwa AF, Kalluvya S, Changalucha J, Fitzgerald DW, Downs JA. Barriers to access reproductive health care for pregnant adolescent girls: a qualitative study in Tanzania. Actapaediatrica. 2015; 104 (12):1291–7. https://doi.org/10.1111/apa.12886 PMID: 25473729

45. Hailemariam A, Haddis F. Factors affecting unmet need for family planning in southern nations, nationalities and peoples region, Ethiopia. Ethiopian journal of health sciences. 2011; 21(2):77–90. https://doi.org/10.4314/ejhs.v21i2.69048 PMID: 22434988

46. Ayele W, Tesfaye H, Gebreyes R, Gebreselassie T. Trends and determinants of unmet need for family planning and programme options, Ethiopia: Further analysis of the 2000, 2005, and 2011 Demographic and Health Surveys. Calverton, Maryland, USA: ICF International; 2013.

47. Wafula SW. Regional differences in unmet need for contraception in Kenya: insights from survey data. BMC Women’s Health. 2015; 15(1):86. https://doi.org/10.1186/s12905-015-0240-z PMID: 2646667029.

48. Teshale AB. Factors associated with unmet need for family planning in Sub-Saharan Africa: A multilevel multinomial logistic regression analysis. PLoS ONE. 2022; 17(2). https://doi.org/10.1371/journal.pone.0263885

49. Wulifan JK, BrennerS, Jahn A. et al. A scoping review on determinants of unmet need for family planning among women of reproductive age in low and middle income countries. BMC Women’s Health. 2015; 16 (2). https://doi.org/10.1186/s12905-015-0281-3

50. Cleland J, Harbison S and Shah IH. 2014. “Unmet need for contraception: Issues and challenges.” Studies in Family Planning 45(2): 105–122

51. UNAIDS. The Global plan to eliminate new HIV infections among children and keeping their mothers alive. 2011; Available from: https://www.unaids.org/sites/default/files/media_asset/JC2774_2015ProgressReport_GlobalPlan_en.pdf.

